# KNOWLEDGE ABOUT SEXUALLY TRANSMITTED DISEASES AMONG THE STUDENTS OF SELECTED MADRASHAS OF DHAKA CITY

**DOI:** 10.1101/2020.06.20.20136515

**Authors:** Tareq Mahmud

## Abstract

**Background:** Sexually transmitted diseases have become a silent epidemic that devastates the life of both men and women. In Bangladesh, there has been no national prevalence data on Sexually transmitted diseases.

**Aims & objectives:** The objectives of the study were to find out the level of knowledge about sexually transmitted diseases among madrasha students.

**Methods:** A cross-sectional study was conducted in four purposively selected madrashas in Dhaka city. A total randomly selected 390 students of class 9 and 10 were included in this study. A semi structured questionnaire was used to collect data and the responses of students were recorded after obtaining verbal consent.

**Results:** Mean age of the respondents was 17 and 33.1% of them were from Science group. 54% respondents had average knowledge, 29.5% had good knowledge. Television/radio was the main source of information for majority (68.6%) of the students and around 40% was informed about STDs from their academic study. About 75% of the respondents mentioned that there was microorganism for all STDs as causative agents. More than 70% respondents mentioned that unprotected sexual intercourse and using same syringes with different persons were the mood of transmission of STDs. Condom as the main protector of STDs was seemed by the most. Type of madrasha and academic group were significantly related (P<0.00) with level of knowledge on STDs.

**Conclusion:** Science group respondents had more academic knowledge than the arts group respondents and respondents who belonged to late adolescent group had more knowledge than early adolescent group.

## Introduction

Since early eighties, the sexually transmitted diseases (STDs) are a great public health concern in the world particularly in developing countries as it enhances the transmission of Human Immune deficiency Virus (HIV). Besides, sexually transmitted diseases can have serious consequences on reproductive health and well-being of both men and women. Both short- and long-term sequelae of untreated Sexually transmitted diseases cause profound biomedical, social and economic impact on individuals and communities. Thus, the control of Sexually transmitted diseases is now recognized as a global priority^1^.Sexually transmitted diseases cause a large proportion of the global burden of ill-health. WHO estimates that over 340 million new cases of four curable STDs (gonorrhea, chlamydia, syphilis and trichomoniasis) occurred in 1999. If viral STDs such as human papilloma virus (HPV), herpes simplex virus (HSV) and human immunodeficiency virus (HIV) infections are included, the number of new cases may be three times higher. Among women, non-sexually transmitted are even more common^2^. Sexually transmitted Diseases (STDs) are those diseases that are contracted mainly through sexual intercourse. They include curable ones like gonorrhea, syphilis, and chlamydia infection as well as incurable but modifiable ones like HIV, herpes simplex, human papilloma virus (HPV), and hepatitis B infections. Sexually transmitted infections (STI) are spread primarily through person-to-person contact, although some of the pathogens that cause it, especially Human immunodeficiency virus (HIV) and syphilis can be transmitted from mother to child during pregnancy and childbirth, and through blood products and tissue transfer^3^.Iatrogenic infections e.g. infection resulting from such unhygienic medical procedures like unsafe abortion, child birth under unhygienic conditions and post-operative infections. STDs may also result from unhygienic health practices such as use of unclean menstrual protection, poor sexual hygiene, vaginal douching, etc. There is a clear distinction between the reproductive tract infections and sexually transmitted diseases. RTIs include all infections of the reproductive tract, whether they are sexually transmitted or not. Bacterial vaginosis and candidiasis, and pelvic inflammatory diseases (PID) caused by iatrogenic infections are considered as reproductive tract infection, but are not sexually transmitted. On the other hand, the pathogens, which are commonly transmitted through sexual contact (HIV, hepatitis B, C, D, etc.), are not always, or sometimes not at all, an infection of the reproductive tract^1^.There are more than 20 types of STDs, including Chlamydia, Genital herpes, Gonorrhea, HIV/AIDS, HPV, Syphilis, Trichomoniasis. Most STDs affect both men and women, but in many cases the health problems they cause can be more severe for women. If a pregnant woman has an STD, it can cause serious health problems for the baby. Antibiotics can treat STDs caused by bacteria, yeast, or parasites. There is no cure for STDs caused by a virus, but medicines can often help with the symptoms and keep the disease under control. Correct usage of latex condoms greatly reduces, but does not completely eliminate, the risk of catching or spreading STDs. The most reliable way to avoid infection is to not have anal, vaginal, or oral sex^4^.Many STDs have no signs or symptoms (asymptomatic). According to WHO, the prevalence of Sexually transmitted diseases is much higher in most developing countries than in developed countries In Bangladesh, little is known about STDs prevalence, social context of their transmission, and of their sequelae. However, a limited number of prevalence studies points to an alarming situation. Almost all of these works are urban based focusing the high-risk group of population^1^. The Madrasha system continued to be attached to the Islamic aspects of the Bangladeshi identity; Further, the neglect of the Madrasha system during the time of colonization created an aura of backwardness about it compared to the modern educational system. This had a strong influence on the negative views of the educated middle class and elites of Bangladesh. They adopted the view, and for the most part continue to believe, that the Madrasha system is a backward one, and that it must be replaced by the modern educational system. However, it is considered as convenient educational institute for mostly rural areas as well as lower middle- and lower-class people of urban areas^17^. Currently, more Madrashas are becoming Alia Madrasha which is incorporating the general education curriculum along with a religious curriculum. Similar to trends among Islamic educational institutions in other Muslim countries, Alia Madrashas offer both religious education and modern general education. The other type of Madrasha, the Qawmi Madrasha, is a non-governmental educational institution; it is a private system of Madrasha education.

## Methodology

A cross sectional study design with quantitative method was conducted with the students of class 9 and 10 in four selected madrashas from Dhaka North and Dhaka South city corporation. From Dhaka south city corporation Darunnazat Siddiqiya Kamil madrasha, alia madrasha which was situated at Shukurshi, under Demra thana of, Madinatul Ulum Madrasha as Qawmi from East dogair, Mohakash school road, sarulia, Demra, Dhaka. From Dhaka north city corporation Quaderia Taiyebia Alia Kamil Madrasa as alia madrasha from Madrasha len, Mohammadpur, and Zamia Rahmania Madrasha, near saat masjid lane and park, Mohammadpur, Dhaka.

Estimated sample size was 384, assuming some loss during interview due to unpredicted reasons, 390 samples were taken.

A semi structured interviewer administered questionnaire was developed and finalized for data collection after pretesting among 10 madrasha students similar to the study population other than the study area (Dogair Darus-Sunnat Fazil Madrasha, located in dogair bazar, demra, Dhaka). After finalization of the questionnaire, data were collected by interviewer administered questionnaire. The questionnaire was consisting of 3 parts. Part 1 was general information of the respondents such as identification number, name, address which was autonomous and maintained confidential. The 2^nd^ part was consisting of socio-demographic information where there were ten questions. Part 3 was the knowledge related part consists of eleven close ended questions. Initially there was an open-ended question for the yes answer of question number 11, but after pretesting it was presented as close ended question and finally another question was source of information related which had the rights to provide multiple response for each respondent.

To find out the level of knowledge of each respondent about STDs, a percentage scale was developed by the researcher. Each correct response scored one mark and no response or wrong response had scored zero mark. Those who scored considered as 0-20% had very poor knowledge, 21-40% had poor knowledge, 41-60% had average knowledge, 61-80% had good knowledge and above 80% had excellent knowledge. Initially all the responses were checked for their completeness, correctness in order to exclude missing or inconsistent data Corrected data were entered into the computer. The data were analyzed by using the Statistical Package for Social Science (SPSS) 16. Collected data were analyzed by descriptive statistics (frequency distribution, Percentage, mean and standard deviation) and Chi-square test to find out the relationship between variables. Statistical significant was set at 95% Confidence Interval.

Approval was obtained from Ethical Review Committee of State University of Bangladesh.Administrative permission was taken from selected Madrasha’s authorities and verbal informed consent was taken from each participant.Privacy and confidentiality were maintained strictly.

## Results

The study included 390 Madrasahs students with a response rate of 100%. Table 1 presents the socio-demographic characteristics of the participants. Their age ranged between 14 and 21 years. 164 respondents (42.1%) were from mid adolescent ages and 226 respondents (57.9%) are were in Late adolescent ages where 129 respondents were from Sciences and 261 respondents were from Arts group. 8 students’ fathers (2.1%) are illiterate while 80 (20.5%) have at least H.S.C level of education. 2 (.5%) of students’ mothers are illiterate and 31 (7.9%) have at least university level of education.

**Table 1.** Sociodemographic characteristics of the participants (n=390)

| **Table-1: Sociodemographic characteristics of the participants (n=390)** |  |  |  |
| --- | --- | --- | --- |
|  | Characteristics | No. | % |
| Age | Mid Adolescent (14 to 17 year) | 164 | 42.1 |
|  | Late Adolescent (18 to 21 year) | 226 | 57.9 |
| Class | 9 | 197 | 50.5 |
|  | 10 | 193 | 49.5 |
| Academic group | Science | 129 | 33 |
|  | Arts | 261 | 67 |
| Father's educational qualification | Illiterate | 8 | 2.1 |
|  | Primary | 28 | 7.2 |
|  | Secondary | 34 | 8.7 |
|  | S.S.C | 98 | 25.1 |
|  | H.S.C | 104 | 26.7 |
|  | Graduation | 80 | 20.5 |
|  | Masters | 26 | 6.7 |
| Mother's educational qualification | Illiterate | 2 | 0.5 |
|  | Primary | 58 | 14.9 |
|  | Secondary | 142 | 36.4 |
|  | S.S.C | 86 | 22.1 |
|  | H.S.C | 71 | 18.2 |
|  | Graduation | 31 | 7.9 |
| Family Type | Nuclear | 331 | 85 |
|  | Joint | 59 | 15 |
| Family Members | ≤ 4 | 155 | 39.7 |
|  | 8-May | 195 | 50 |
|  | More than 8 | 40 | 10.3 |
|  | Minimum 2, Maximum 12 |  |  |

**Figure 1:**
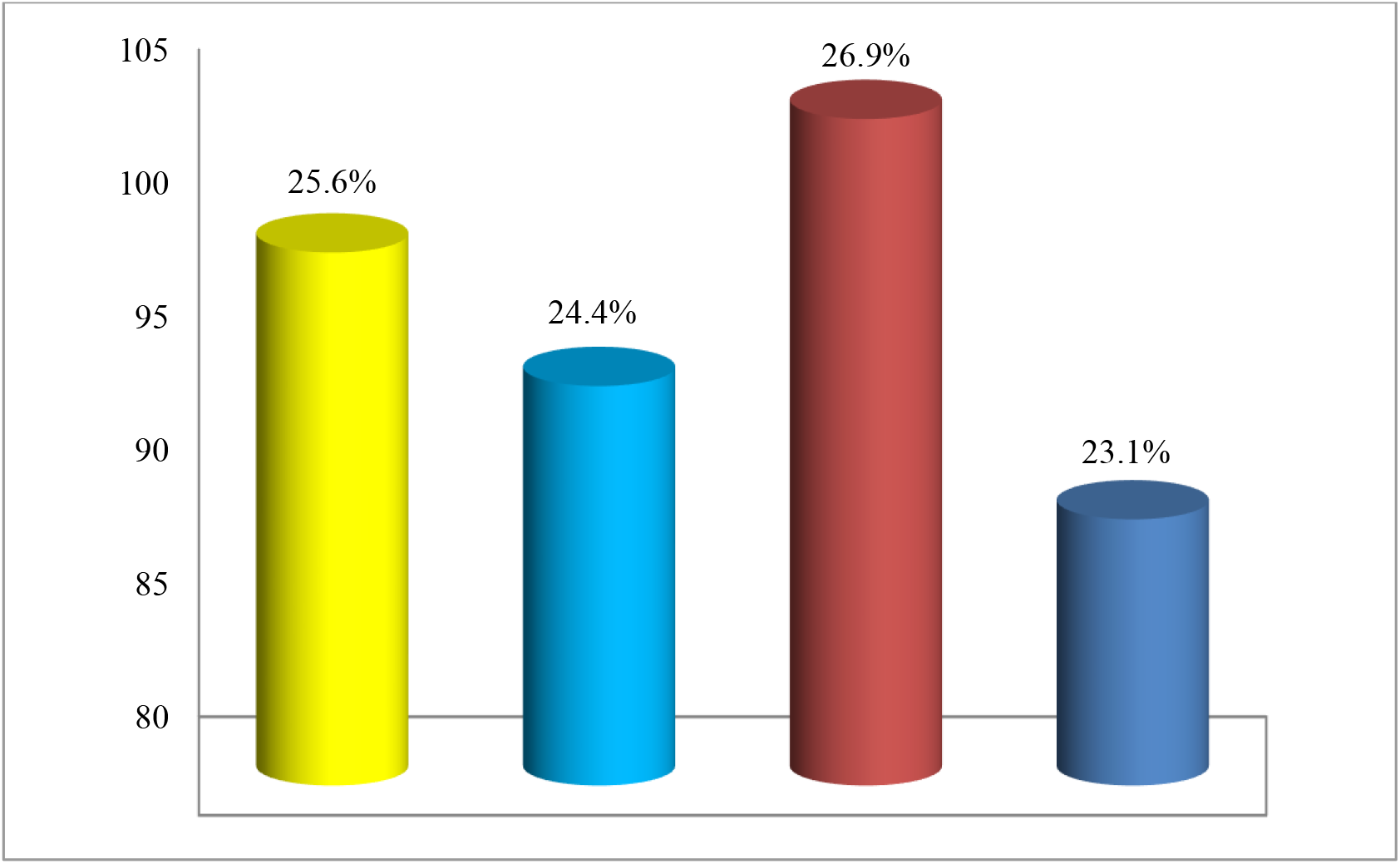
Distribution of respondents according to Madrasha type: (n= 390)

This figure illustrates the distribution of respondent according to Madrasha type. It narrates the percentage of respondent study in madrasha 25.6% respondent was from Alia Madrasha of South City Corporation and 26.9% from North City Corporation. 24.4% respondent from Qawmi Madrasha of South City Corporation and 23.1% from North City Corporation.

**Table 2.** Participant’s knowledge regarding different aspects of STDs (n=390)

| Question | Right answer |  |
| --- | --- | --- |
|  | Frequency | % |
| **Mode of transmission** |  |  |
| STDs could be transmitted from use of same syringes with different person | 281 | 72 |
| STDs could be transmitted by sharing the same plate with infected person | 281 | 72 |
| STDs could be transmitted by unprotected sexual intercourse | 292 | 75 |
| STDs could be transmitted from sharing the same toilet with an infected person | 316 | 81 |
| STDs could be transmitted by exposure to cough and sneeze from infected person | 248 | 64 |
| STDs could be transmitted from only one sexual contact | 209 | 53.6 |
| STDs could be transmitted through ways other than sexual contact. | 169 | 43.4 |
| STDs infected mother could transmit a sexual disease to her newborn during labor | 317 | 81.3 |
| STDs infected father could transmit a sexual disease to his newborn if mother is not infected | 246 | 63 |
| **Preventive measures** |  |  |
| Is condom could protect from STDs 100% | 322 | 82.6 |
| Is using of Oral contraceptives could decrease the risk of STDs among women | 158 | 40 |
| Is there any vaccination for STDs protection | 160 | 41 |
| **Causative agents** |  |  |
| HIV is the main causative agents | 87 | 22.3 |
| Is there any common microorganism of STDs | 291 | 74.6 |

Table shows that in majority of the respondents 72% (n= 281) answered STDs are transmitted from use of same syringes with different person and Sharing the same plate with infected person, 75% (n= 292) answered STDs are transmitted by unprotected sexual intercourse. 81.3% (n= 317) answered that STDs infected mother could transmit a sexual disease to her newborn during labor and 18.7% (n=73) answered STDs infected mother could not transmit a sexual disease to her newborn during labor. 322 students (82.6%) know that condom can prevent STDs, while they also had some misconception and 291 students (74.6%) mentioned that there is common microorganism for STDs.

**Figure 2:**
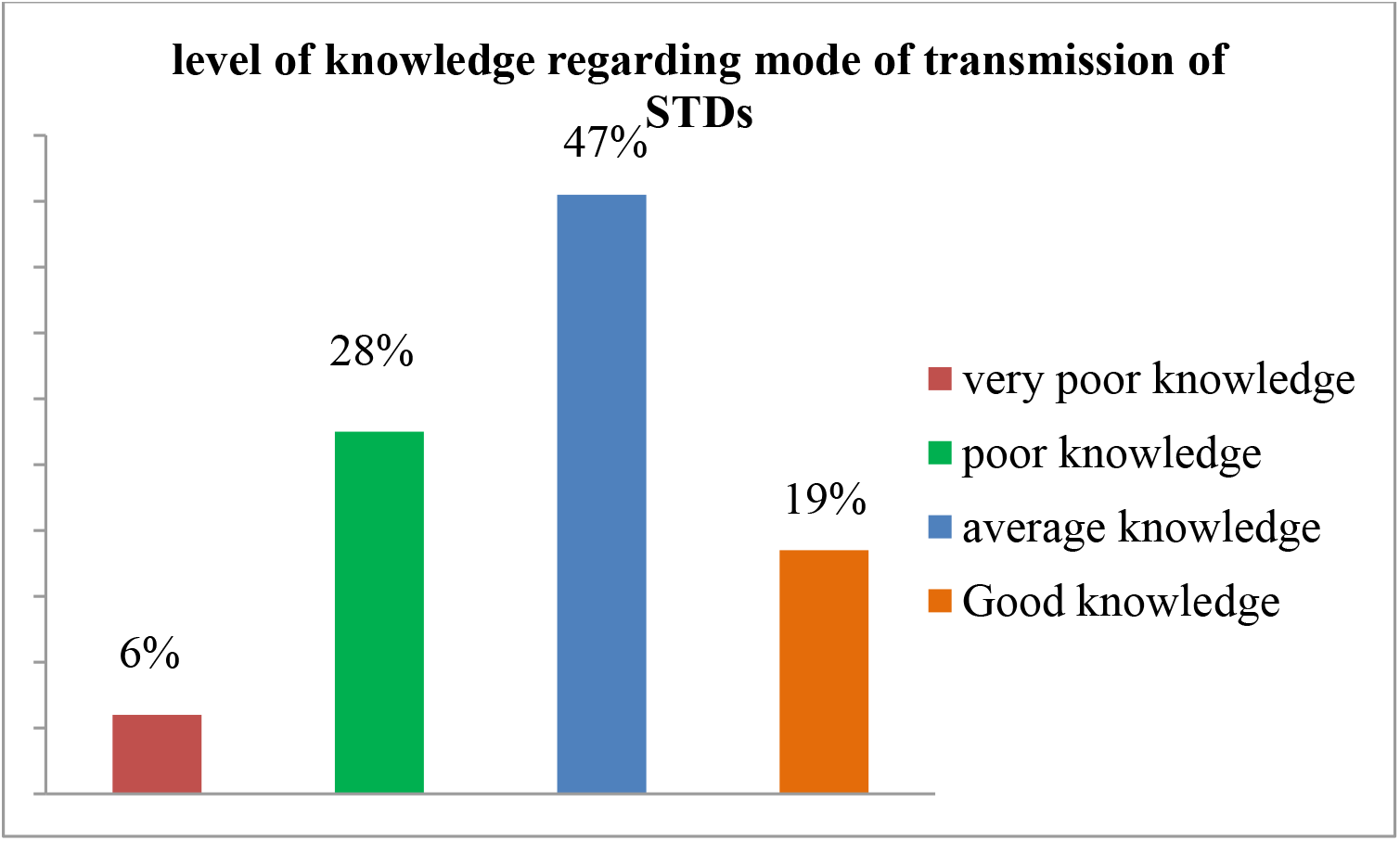
Distribution of respondents according to level of knowledge regarding mode of transmission of STDs (n= 390)

Figure illustrates that respondents 47% (n= 182) had average knowledge, 19% (n= 74) had good knowledge, 28% (n= 110) had poor knowledge and 6% (n= 24) had very poor knowledge regarding mode of transmission of STDs.

**Table 3:** Distribution of respondents according to level of knowledge regarding preventive measures and causative agents of STDs.

| Variables | Level of knowledge |  |  |  |
| --- | --- | --- | --- | --- |
|  | Poor | Average | Good | Excellent |
| **Preventive measures** |  |  |  |  |
| Frequency | 204 | 112 | 74 | 10 |
| % | 50 | 29 | 19 | 2 |
| **Causative agents** |  |  |  |  |
| Frequency | 62 | 278 | 50 | 0 |
| % | 16 | 71 | 13 | 0 |

Table narrates that respondents 71% (n= 278) had average knowledge, 13% (n= 50) had good knowledge and 16% (n= 62) had poor knowledge regarding causative agents of STDs and respondents 29% (n= 112) had average knowledge, 19% (n= 74) had good knowledge, 50% (n=204) had poor knowledge and 2% (n= 10) had excellent knowledge regarding preventive measures of STDs.

**Table 4:** Distribution of respondents according to Source of information (n= 390)

| **Source of information** | **Frequency (n)** | **Percentage (%)** |
| --- | --- | --- |
| TV/radio | 258 | 68.6% |
| Newspaper | 157 | 41.8% |
| Parents/ Relatives/ Neighbors | 46 | 12.2% |
| Friends/ teachers | 230 | 61.2% |
| Academic knowledge | 153 | 40.7% |
| Public talks/seminars | 68 | 18.1% |
| Billboards/posters | 150 | 39.9% |
| No response | 14 | 3.6% |

**\*Multiple responses**

Table narrates that majority of the respondents 68.6% (n= 258) source of information was TV/radio, followed by 41.8% (n= 157) responded their source of information was newspaper, and 12.2% (n=46) respondents respond that they heard from their Parents/ Relatives/ Neighbors, 40.7% (n= 153) respond their source of information was their academic knowledge, 18.1% (n=68) respondents respond that they heard from public talks/seminars, 39.9% (n= 150) respond their source of information was billboards/posters and 3.6% (n= 14) out of 390 did not response for any options.

**Table 5:** Association between knowledge score and different variables of the students (n=390)

| Variables | Level of knowledge |  |  |  | P Value |
| --- | --- | --- | --- | --- | --- |
|  | Poor | Average | Good | Excellent |  |
| Types of Madrasahs |  |  |  |  | 0 |
| Alia | 12 (6%) | 53(26%) | 89 (43%) | 51 (29%) |  |
| Qawmi | 21 (10.52) | 138 (73.68%) | 17 (10.52%) | 9 (5.26%) |  |
| Academic group |  |  |  |  | 0 |
| Science | 2 (16.66%) | 24 (11.3%) | 60 (52.2%) | 43 (84.3%) |  |
| Arts | 10 (83.33%) | 188 (88.7%) | 55 (47.8%) | 8 (15.7%) |  |
| Age |  |  |  |  | 0 |
| Mid Adolescent | 8 (60%) | 74 (35%) | 57 (49.6%) | 25 (49%) |  |
| Late Adolescent | 4 (40%) | 138 (65%) | 58 (50.4%) | 26 (51%) |  |

Table shows that Alia madrasahs students level k knowledge is better than the Qawmi madrasahs students. Where science groups students have more knowledge than the arts group students. Age is also a associated factors with knowledge.

**Figure 3:**
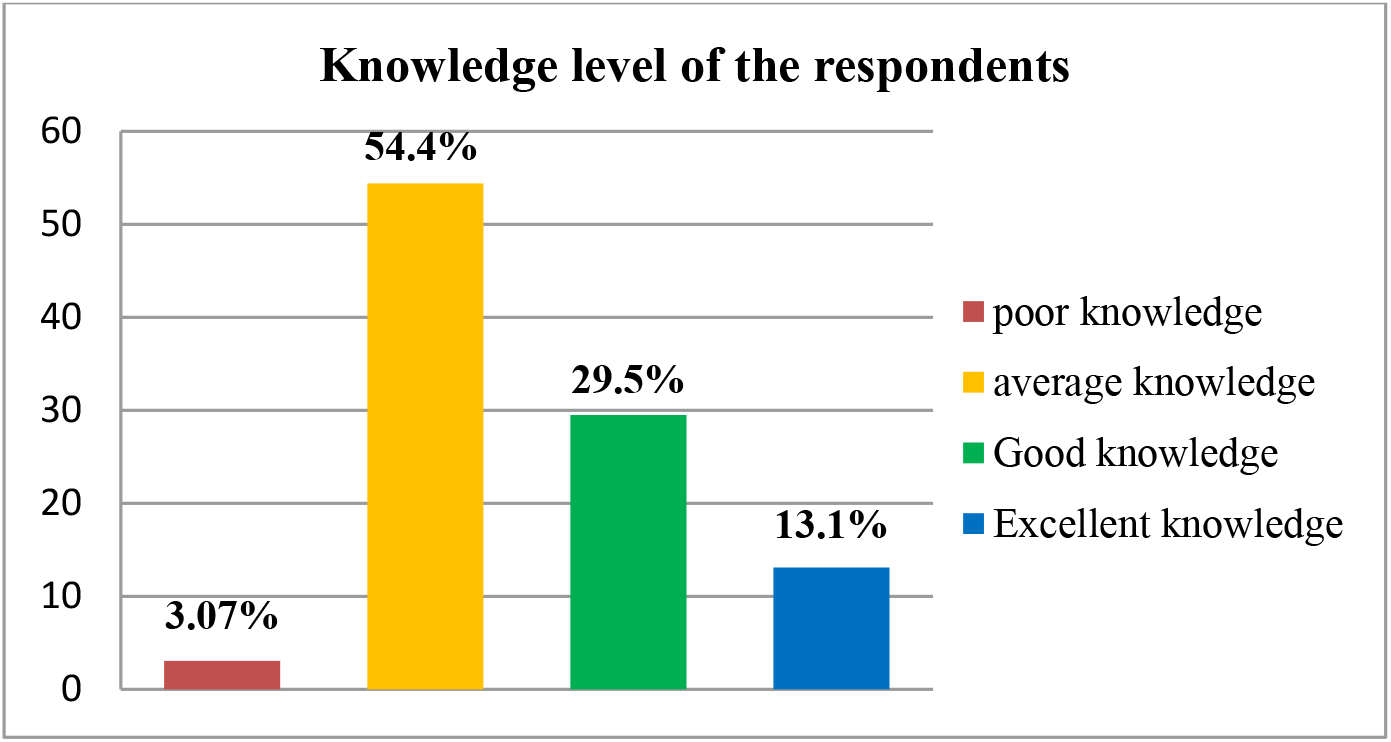
Distribution of respondents according to overall level of knowledge (n= 390)

Figure illustrates that respondents 54.35% (n= 212) had average knowledge, 29.5% (n= 115) had good knowledge, 13.1% (n= 51) excellent knowledge and 3.07% (n= 12) had poor knowledge.

## Discussion

.A large number of respondents (71.2%) had mentioned that sexually transmitted disease transmitted by use of same syringes with different person followed by (77.2%) had mentioned the way as unprotected sexual intercourse. About (74.6%) mentioned that there was common microorganism for all STDs. Around (37.6%) had misconception about transmission as exposure to cough and sneeze from infected persons, (28.8%) said sharing the same plate with infected person as a cause, whereas (19.6%) responded sharing the same toilet with an infected person. A study was conducted in Nigeria which indicates that (87.6%) mentioned the way as unprotected sexual intercourse, where (22.0%) had misconception about cough and sneeze,(16.1%) had misconception about sharing toilets and (12.2%) had misconception about sharing same plates with infected person ^15^. Also, regarding transmission of STDs (81.2%) said that an infected mother could transmit a sexual disease to her newborn during labor and (63.1%) said that an infected father could transmit a sexual disease to his newborn if mother is not infected which was a misconception. Whereas the study conducted in Saudi Arabia revealed that (36.8%) an infected mother could transmit a sexual disease to her newborn during labor and (45.0%) said infected father could transmit a sexual disease to his newborn if mother is not infected^18^. This indicates clearly that study respondents had better knowledge about transmission by mother and less knowledge of transmission by father. Respondent’s responded regarding preventive measures against sexually transmitted diseases (93.1%) had said that condom could protect from STDs 100%. Also, they had some misconception that oral contraceptives could decrease the risk of STDs among women (67.1%) and there is vaccination for STDs protection (66.5%). A study conducted in Saudi Arabia showed that (47.0%) said condom could protect from STDs. And also, they had misconception that oral contraceptives could decrease the risk of STDs among women (54.3%) and vaccination for STDs protection (38.0%)^8^. Compared to both study respondents of this study had less knowledge about preventive measures of STDs. In this study 54.35% respondents had average knowledge, 29.48% had good knowledge, 13.1% excellent knowledge, and 3.07% had poor knowledge. Comparing to age of the respondents in mid adolescent only 6 respondents had poor knowledge, (34.9%) had average knowledge, (49.6%) had good knowledge, and (49.0%) had excellent knowledge whereas in late adolescent 4 respondents had poor knowledge, (65.1%) had average knowledge, (50.4%) had good knowledge, and (51.0%) had excellent knowledge. Comparing to the type of madrashas at alia madrasha (43.41%) respondents had good knowledge, (5.85%) had poor knowledge, (25.85%) had average knowledge and around 25% had excellent knowledge. At Qawmi madrasha majority of respondents (74.60%) had average knowledge, (11.35%) had poor knowledge, (9.2%) had good knowledge and only (4.86%) had excellent knowledge. Comparing to the group of the respondents and their academic knowledge, in science group n=125 (81.7%) had academic knowledge and n= 4 (1.7%) they did not mention about their academic knowledge, whereas in arts group n= 233 (98.3%) did not had academic knowledge and n= 28 (18.3%) had academic knowledge. Comparing to the group of the respondents (66.9%) were from arts group and majority of the respondents (72.03%) of them had average knowledge, (21.07%) had good knowledge, (3.83%) had poor knowledge and only (3.06%) had excellent knowledge whereas in science groups 24 (18.6%) had average knowledge, (46.51%) had good knowledge, (1.55%) had poor knowledge and (33.33%) had excellent knowledge compared to arts group. So, it clearly indicates that science group respondents had more knowledge than arts group. Educational activities at schools should be increased in order to better inform the students of these problems as it is very important to offer young people better and more correct information about STDs and HIV/AIDS ^37-40^. Most of the respondents (68.6%) mentioned that source of information was TV/radio, followed by 41.8% responded their source of information was newspaper, and 12.2% respondents respond that they heard from their Parents/ Relatives/ Neighbors, 40.7% respond their source of information was their academic knowledge, 18.1% respondents respond that they heard from public talks/seminars and 39.9% respond their source of information was billboards/posters. Previous study conducted in Saudi Arabia showed that (27.7%) had known from TV, (15.3%) had known from books/ academic knowledge, (35.0%) had known from friend/ relatives and (29.9%) had known from internet^22^.So, in comparison more respondents had known from TV, academic knowledge rather than parents/ relatives/ friend from the previous study. People were found to be more informed about HIV/AIDs than the other STDs due to media coverage and publicity ^41^.

## Conclusion

Around 54% of the respondents had average knowledge and near 30% had good knowledge about STDs. About 75% of the respondents mentioned that there was same microorganism for all STDs as causative agents. More than 70% respondents mentioned that unprotected sexual intercourse and using same syringes with different persons were the mood of transmission of STDs. Condom as the main protector of STDs was seemed by the most (82.6%) of them. Television/radio was the main source of information for majority (68.6%) of the students and around 40% was informed about STDs from their academic study. Age, type of madrasha and academic group were significantly related (P<0.00) with level of knowledge on STDs. Science group respondents had more academic knowledge than the arts group respondents. Respondents of Alia madrasha had comparatively better knowledge than the respondents of Qawmi madrasha. Respondents who belonged to late adolescent group had more knowledge than early adolescent group.

## Recommendations

This study was conducted only in four selected madrashas of Dhaka city which might not reflect the actual result representing the level of knowledge of the population all over the country. Therefore, further large-scale study is recommended.

Though only two fifth students were informed about STDs from their academic study, more information specifically modes of transmission, symptoms, causes and ways of prevention of STDs should be included in madrasha curriculum mostly in the Qawmi madrasha and for arts groups of Alia madrasha.

## Data Availability

All data will be available for viewing, all data can be stored.

## Acknowledgement

I would like to express my deepest regards and heartfelt gratitude to my respected supervisor Nuhad Raisa Seoty, Assistant Professor, Department of Public Health, State University of Bangladesh for proper guidance, constructive criticism and whole hearted cooperation during my research work.Special gratitude to Prof. Dr. Nawzia Yasmin, Professor and Head, department of Public Health, State University of Bangladesh for their continuous and patience guidance.

## Notes

### Competing Interest Statement

The authors have declared no competing interest.

### Funding Statement

No funding. Author's own interest

### Author Declarations

A DISSERTATION SUBMITTED IN PARTIAL FULFILLMENT OF THE REQUIREMENT FOR THE DEGREE OF MASTER OF PUBLIC HEALTH (MPH), ALL PERMISSION GRANTED FROM ETHICAL COMMITTEE OF SUB

